# Patient Safety of Remote Primary Care: A Qualitative Study Assessing Risks and Mitigation and Prevention Strategies

**DOI:** 10.1101/2024.01.30.24301946

**Authors:** Olivia Lounsbury, Edmond Li, Tetiana Lunova, Jackie van Dael, Niki O’Brien, Ahmed Alboksmaty, Alay Rangel-Cristales, Ara Darzi, Ana Luisa Neves

## Abstract

**Background:** While virtual care delivery has numerous advantages, it can also introduce safety risks and unintended consequences. Considering that it has become an integral part of today’s healthcare service, uncovering its unintended consequences is imperative to ensure patient safety.

**Objectives:** This study aimed to identify patient safety risks associated with virtual primary care, as well as strategies to mitigate these risks based on the perspectives of patients and healthcare providers.

**Methods:** Three focus groups were conducted followed by semi-structured interviews with patients, carers and healthcare providers working in primary care. Transcripts were systematically reviewed, and thematic analysis was performed by two independent researchers.

**Results:** A total of 42 participants took part in the study. Three main areas for patient safety risks associated with virtual primary care were identified, including suboptimal clinical decision-making, negative impact on patients’ access to care, and worsening the workforce crisis. Strategies to mitigate these risks included providing information for patients, training triage personnel, making technical support available, standardising guidelines, setting up systems for feedback, improving continuity of care, communication, and safety netting.

**Conclusions:** Patients and providers now have a heightened awareness of the strengths and pitfalls of virtual care due to their increased familiarity with the use of virtual care technologies. Existing policies need to be updated and new ones devised to minimise risks associated with virtual care and support patient and provider workflows.

**Public Interest Summary:** The COVID-19 pandemic galvanized an emergent necessity to deliver care virtually in order to reduce disease transmission. However, given the urgency of the crisis, virtual care was being delivered with minimal protective measures for safety.

This study examines the lived experiences of both patients and providers around virtual care use in England. Potential risks of virtual care delivery, and strategies to mitigate these risks, are identified from both perspectives. The risks identified vary from the technological learning curve to the challenges associated with modified patient-provider communication. The potential solutions identified range from strategies to improve micro-level patient-provider interactions to larger-scale system changes to improve the continuity of care.

Support for patients and providers alike should be allocated to alleviate unnecessary burdens associated with virtual care. Ensuring patient safety necessitates seamless coordination and interoperability between virtual and in-person healthcare to maintain harmony between the two modes of healthcare delivery.

## Introduction

Medical care has been provided via telemedicine for several decades and was adopted initially to reduce healthcare costs, improve quality, and expand access to care (1, 2). Despite years of investment and innovation, it was not until the start of the COVID-19 pandemic that virtual care became widely implemented. The rapid spread of the SARS-CoV-2 virus abruptly forced patients, providers, and healthcare systems to make significant adjustments to their routine practices for delivering and receiving healthcare (3). To adapt to the emerging challenges and reduce COVID-19 transmission, virtual services, including phone, video, or online consultations, quickly became a major route of primary care delivery (4, 5).

The COVID-19 pandemic exposed the potential advantages of virtual care, with some studies indicating a possible role for more efficient patient triage, reduction of delays in diagnosis and treatment, lower travel costs, and improved physician work flexibility (3, 6, 7). The shift from in-person to virtual consultations has minimised virus transmission while maintaining a comparable quality of care and relatively high patient satisfaction levels (8). Virtual care has expanded access for some groups, but it has been acknowledged that access can be difficult for those without stable internet, or with hearing or visual impairments (9, 10). Virtual care has also been associated with healthcare cost reduction and lower hospital admission rates in certain population groups, such as older persons (11).

While this means of care delivery can potentially address several challenges associated with face-to-face care delivery, particularly for emergency situations like the COVID-19 pandemic, it can also introduce safety risks and unintended consequences. Research into patient perspectives suggests patients have lower confidence in the safety of virtual care - particularly virtual consultations -, stemming from concerns about the clinician’s inability to read their body language, difficulties in communicating effectively, and fewer opportunities for patients to raise issues (12, 13). Some evidence also suggests that virtual consultations, particularly via telephone, are helpful in addressing minor conditions but may increase the likelihood of missing rare but serious conditions (13). Given the abrupt introduction of virtual care delivery due to the pandemic, its safety risks are not largely understood.

Because virtual care has become an integral part of today’s healthcare service, uncovering its risks and exploring potential strategies are imperative for safe, high-quality care delivery. There is limited knowledge of patients’ and physicians’ perceptions of safety risks associated with virtual primary care. As frontline users, both patients and service providers have the potential to identify different safety issues; it is therefore vital to consider their views to comprehensively define risks and mitigation strategies (14). The aim of the current research is to identify patient safety risks associated with virtual primary care, as well as strategies to mitigate these risks, based on the perspectives of patients and healthcare providers.

## Methods

### Overview of the Methods Used

A qualitative approach consisting of online focus groups and semi-structured interviews was adopted. The team includes researchers trained in qualitative research with backgrounds in clinical medicine, public health, and patient safety. The study reporting was performed in accordance with the Consolidated Criteria for Reporting Qualitative (COREQ) checklist.

### Study Population

Study participants included patients and healthcare workers working in a primary care setting (*e.g.,* general practitioners (GP), community nurses, pharmacists). Patients were included if aged 18 years or older and fluent in English.

### Recruitment

Patient participants were recruited via charity newsletters (e.*g.*, Open Age), social media (i.e., LinkedIn, Twitter), online platforms (*e.g.*, VOICE UK and People in Research), and the Research Partners Group at the Patient Safety Translational Research Centre at Imperial College London. Participants for the focus groups and interviews were recruited through the researchers’ professional networks, research networks and social media (i.e., LinkedIn, Twitter) using a convenience sampling approach. All participants were given background on the study and informed of its purpose.

### Data Collection

Three, 60-minute focus groups were conducted between September and November 2021, with a total of 16 participants (eight patients, four GPs, two nurses, one pharmacist, and one preferred not to answer). Each focus group included five to six participants. Two researchers were present in each of the focus groups (JvD, EL), one facilitating the discussion while the other taking field notes. A total of 26 interviews were completed by two researchers (OL, EL) between February and May 2022, including eight general practitioners, one healthcare student, 11 patients and carers (six preferred not to answer). Interviews lasted between 30-60 minutes. Participants did not have previous relationships with the interviewers.

Semi-structured topic guides were used in both the focus groups and interviews (**Appendix 1 and 2**). Focus groups and interviews were conducted in English using remote conference software (*i.e.,* Zoom), recorded and transcribed verbatim. No participant review of the transcripts was performed, and no follow-up focus groups or interviews were conducted.

### Data Analysis

Thematic analysis was performed in which transcripts were systematically reviewed and coded by two independent researchers (15). The coding was both deductive and inductive in its approach and involved an iterative exploration of the data, with initial themes discussed and revised within the interdisciplinary research team. Data saturation was determined through discussion with researchers involved in coding once no new themes emerged.

## Results

### Participant Characteristics

A total of 42 participants were included (focus groups, n= 16; interviews, n=26). One individual participated in both the interview and focus group. A full description of the participants is provided in Table 1. Participant identifiers used alongside included quotes are only known to members of the research team and do not correspond with any other identifiers or roles outside of the study.

**Table 1.**
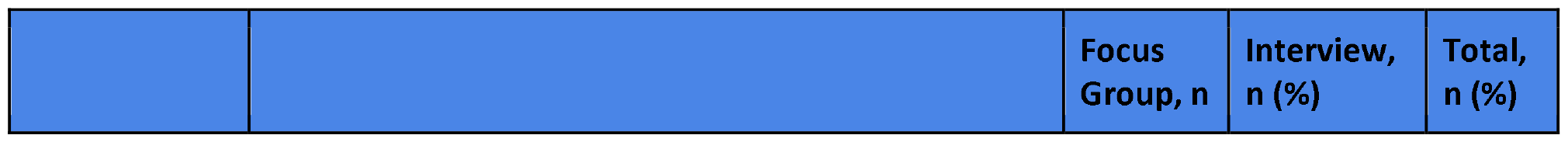
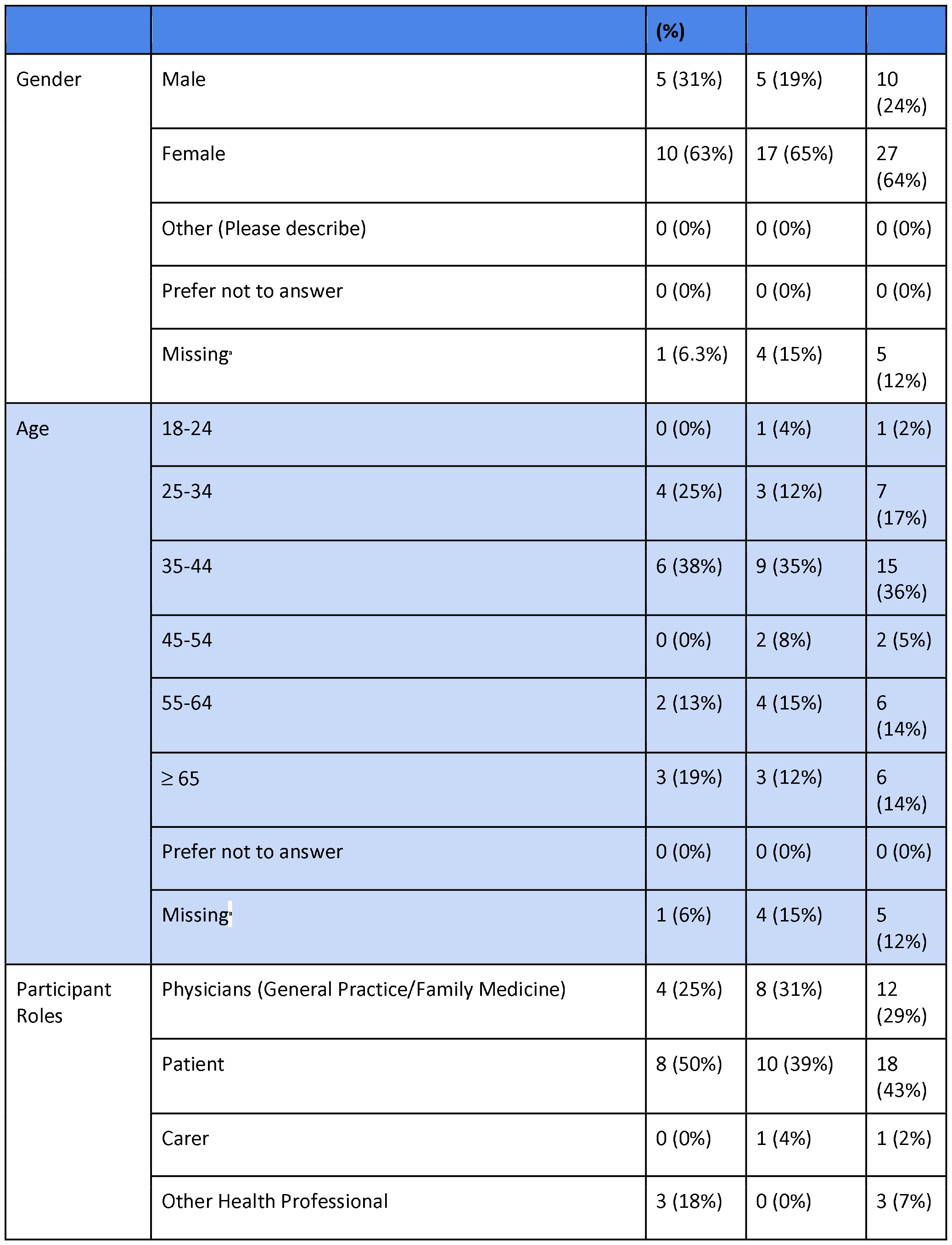
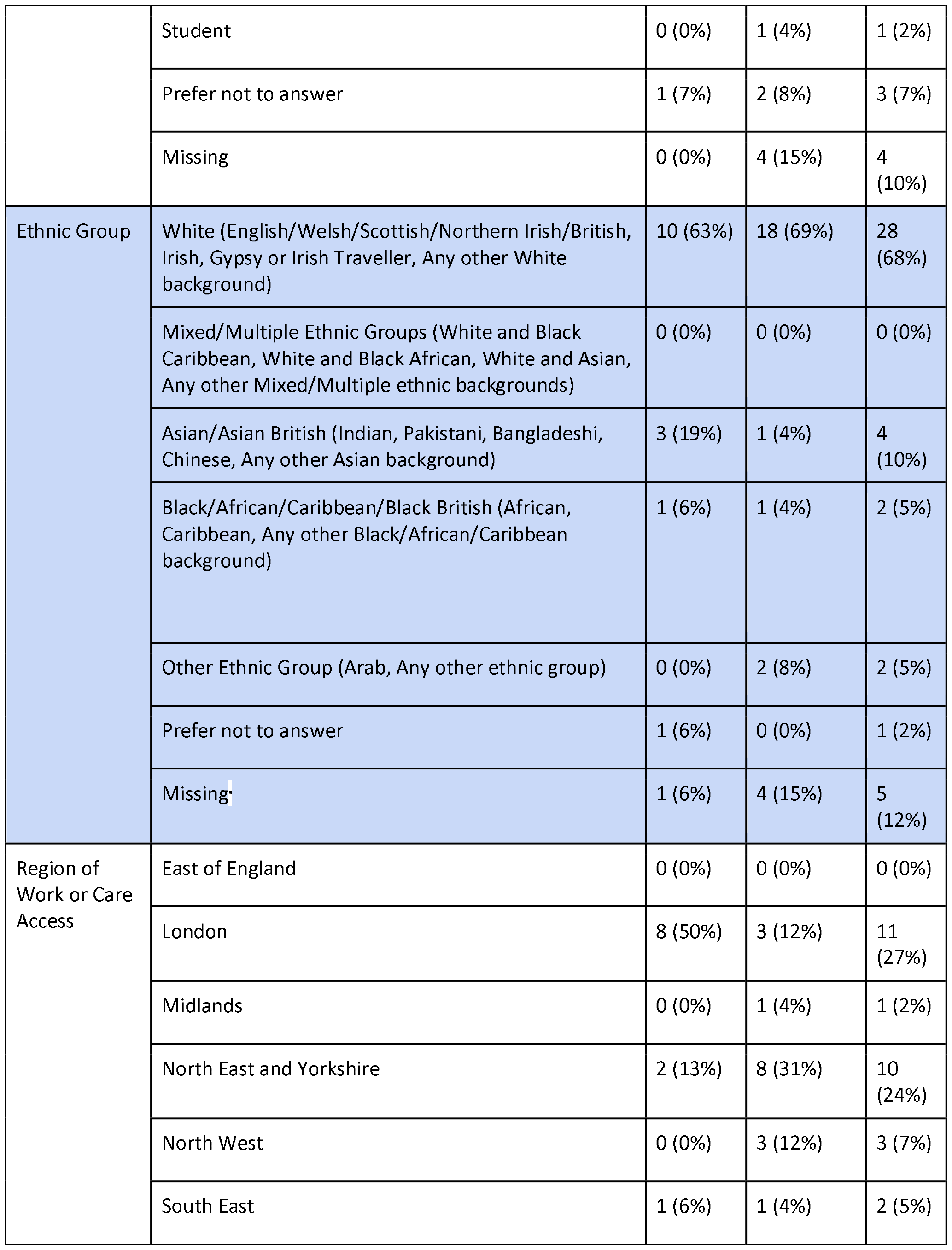
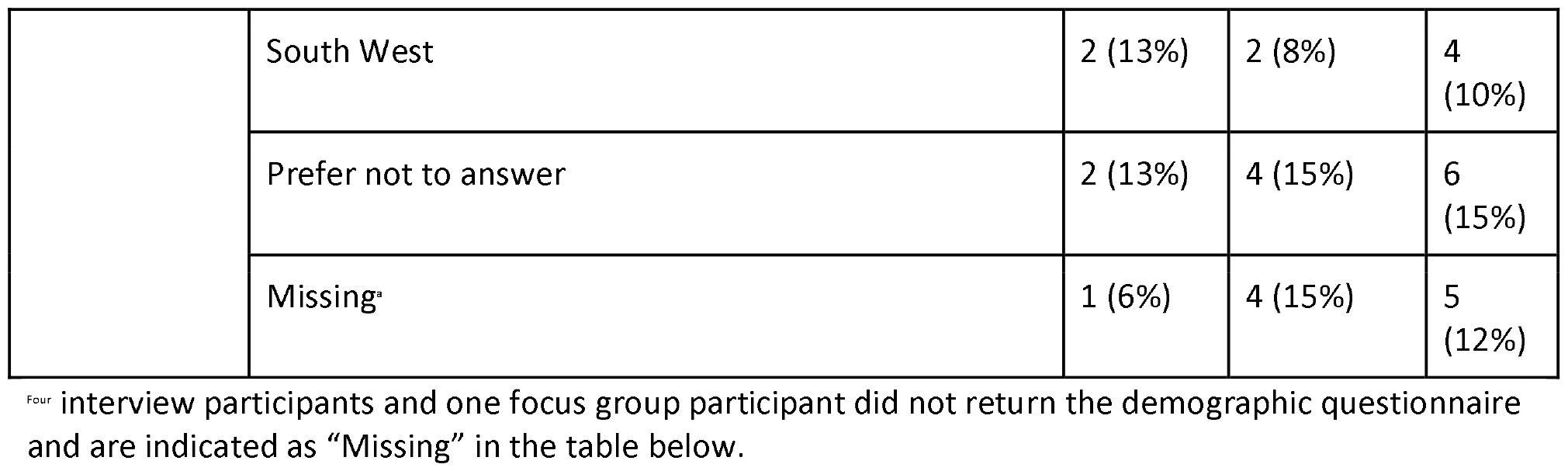
Characteristics of study participants.

|  |  | Focus Group, *n* | Interview, *n* (%) | Total, *n* (%) |
| --- | --- | --- | --- | --- |

|  |  | (%) |  |  |
| --- | --- | --- | --- | --- |
| Gender | Male | 5 (31%) | 5 (19%) | 10 (24%) |
|  | Female | 10 (63%) | 17 (65%) | 27 (64%) |
|  | Other (Please describe) | 0 (0%) | 0 (0%) | 0 (0%) |
|  | Prefer not to answer | 0 (0%) | 0 (0%) | 0 (0%) |
|  | Missing<sup>a</sup> | 1 (6.3%) | 4 (15%) | 5 (12%) |
| Age | 18-24 | 0 (0%) | 1 (4%) | 1 (2%) |
|  | 25-34 | 4 (25%) | 3 (12%) | 7 (17%) |
|  | 35-44 | 6 (38%) | 9 (35%) | 15 (36%) |
|  | 45-54 | 0 (0%) | 2 (8%) | 2 (5%) |
|  | 55-64 | 2 (13%) | 4 (15%) | 6 (14%) |
|  | ≥ 65 | 3 (19%) | 3 (12%) | 6 (14%) |
|  | Prefer not to answer | 0 (0%) | 0 (0%) | 0 (0%) |
|  | Missing<sup>a</sup> | 1 (6%) | 4 (15%) | 5 (12%) |
| Participant Roles | Physicians (General Practice/Family Medicine) | 4 (25%) | 8 (31%) | 12 (29%) |
|  | Patient | 8 (50%) | 10 (39%) | 18 (43%) |
|  | Carer | 0 (0%) | 1 (4%) | 1 (2%) |
|  | Other Health Professional | 3 (18%) | 0 (0%) | 3 (7%) |
|  | Student | 0 (0%) | 1 (4%) | 1 (2%) |
|  | Prefer not to answer | 1 (7%) | 2 (8%) | 3 (7%) |
|  | Missing | 0 (0%) | 4 (15%) | 4 (10%) |
| Ethnic Group | White (English/Welsh/Scottish/Northern Irish/British, Irish, Gypsy or Irish Traveller, Any other White background) | 10 (63%) | 18 (69%) | 28 (68%) |
|  | Mixed/Multiple Ethnic Groups (White and Black Caribbean, White and Black African, White and Asian, Any other Mixed/Multiple ethnic backgrounds) | 0 (0%) | 0 (0%) | 0 (0%) |
|  | Asian/Asian British (Indian, Pakistani, Bangladeshi, Chinese, Any other Asian background) | 3 (19%) | 1 (4%) | 4 (10%) |
|  | Black/African/Caribbean/Black British (African, Caribbean, Any other Black/African/Caribbean background) | 1 (6%) | 1 (4%) | 2 (5%) |
|  | Other Ethnic Group (Arab, Any other ethnic group) | 0 (0%) | 2 (8%) | 2 (5%) |
|  | Prefer not to answer | 1 (6%) | 0 (0%) | 1 (2%) |
|  | Missing | 1 (6%) | 4 (15%) | 5 (12%) |
| Region of Work or Care Access | East of England | 0 (0%) | 0 (0%) | 0 (0%) |
|  | London | 8 (50%) | 3 (12%) | 11 (27%) |
|  | Midlands | 0 (0%) | 1 (4%) | 1 (2%) |
|  | North East and Yorkshire | 2 (13%) | 8 (31%) | 10 (24%) |
|  | North West | 0 (0%) | 3 (12%) | 3 (7%) |
|  | South East | 1 (6%) | 1 (4%) | 2 (5%) |
|  | South West | 2 (13%) | 2 (8%) | 4 (10%) |
|  | Prefer not to answer | 2 (13%) | 4 (15%) | 6 (15%) |
|  | Missing<sup>a</sup> | 1 (6%) | 4 (15%) | 5 (12%) |

<sup>a</sup> Four interview participants and one focus group participant did not return the demographic questionnaire and are indicated as “Missing” in the table below.

### Patient Safety Risks Associated with Virtual Primary Care

Both patients and GPs indicated that there were a variety of technologies used in the delivery of virtual primary care, including telephone, video, email, chats, mobile apps, and online forms. Three main themes with relevant sub-themes were developed to describe potential patient safety risks associated with the use of virtual primary care.

### a. Suboptimal Clinical Decision Making

Suboptimal clinical decision-making may be a result of limited or misinterpreted patient-provided information, lack of formal guidance, or more defensive clinical practice.

#### Limited or misinterpreted patient provided information

Participants noted the significantly greater need to rely on patient-provided information in virtual care environments, which may be inadequate for a comprehensive and accurate clinical assessment, given the patient’s limited knowledge about which signs are relevant to report. Patients voiced concerns about their ability to assess and report on their signs, particularly related to clinical characteristics perceived as ‘subjective’ (i.e., swelling, redness). Participants also noted that those with mental health conditions may minimise the severity of their symptoms; there were concerns about worsening mental health conditions if a provider is not able to detect subtle signs associated with mental health challenges.

#### Lack of formal guidance

The lack of formal guidance was largely highlighted by providers, including concerns of not having structured guidance for remote provision of care, or the need to improve the limited guidance available on when and how to perform virtual consultation, and when should one escalate to in-person care.

#### Defensive medicine

Providers stated that there was an increased tendency to practise defensive medicine in virtual care. As a result, providers employed strategies such as ordering additional tests, or changing prescribing practices as mechanisms for risk mitigation, which in themselves introduce unnecessary risks for harm (e.g., overprescribing antibiotics). Some healthcare workers described this as a way of minimising a sense of uncertainty that came with not performing physical examinations in person. Some participants highlighted positive adaptations to this uncertainty, including increased safety netting.

### b. Negative Impact on Patients’ Access to Care

Participants identified a number of ways in which virtual care compromised access to care, in particular for already disadvantaged groups.

#### Delays in care

Patients experienced a sense of feeling overwhelmed by having to complete complicated online booking forms or feeling like they were inappropriately advised by a reception at the booking stage (*e.g.,* inappropriate modality of appointment). Delays in care were often noted to be the result of serial, stepwise escalations in care, until an eventual face-to-face appointment, which some providers referred to as “delaying the inevitable”.

#### Poor continuity

Patients perceived minimal continuity with the same provider when receiving care virtually, which contributed to inadequate monitoring practices. Participants highlighted that longitudinal records are more difficult to compile through virtual care visits because many of the assessments providers would conduct, such as gait assessments, are largely impossible virtually.

#### Digital exclusion

Many participants indicated that a significant safety risk when using virtual care technologies is digital exclusion, which could be the result of the lack of access to the technology or stable, high-quality internet, or inability to use the technology itself (e.g., due to low digital literacy or visual/hearing impairments). For some individuals, this could be further compounded by additional challenges such as language barriers and reliance on translation by their informal carers, thus posing privacy issues, especially for sensitive topics (e.g., mental health).

#### Patients’ use of alternative routes/services to access health care

Patients who are unable to access virtual care may be more likely to ignore the health condition until severe deterioration occurred; others noted that some patients will access healthcare in other ways, such as via Accident & Emergency (A&E) services.

### c. Worsening the Workforce Crisis

Participants indicated that the use of virtual care may worsen the existing workforce crisis, particularly via digital fatigue and decreased worker satisfaction.

#### Digital fatigue

The inclusion of virtual care in practice was perceived by providers as ‘another thing to monitor’, a concern that was exacerbated by their past experience with digital systems that did not support the coordination of information they expected.

#### Decreased worker satisfaction

The introduction of virtual care has, in some ways, decreased satisfaction with the professional, according to provider participants. Some noted that offering virtual care has given the impression that providers are not keen to provide care for patients; they felt this was damaging to their morale. Similarly, providers were sometimes dissatisfied, or did not find value in the tasks associated with the provision of virtual care.

##### Textbox 1

**Patient Safety Risks Associated with Virtual Primary Care**

###### Theme 1. Suboptimal Clinical Decision-Making

*Subtheme 1.1.1: Limited or misinterpreted patient provided information*

- “I had to try and explain what my foot was looking like on one issue, which is why in the end I had to go and see the consultant. And actually, it was very difficult. And he was saying, Well, how swollen is it? And I was looking at it thinking I don’t know if it’s very swollen or minimally swollen…In my mind it’s very swollen, but is it? It would have been much, much easier by video to have been able to actually show it… [FG1 P4]
- “A couple of my friends with severe mental health problems will minimise everything and not explain stuff properly and if the healthcare professional isn’t able to pick up on any non-verbal cues then they don’t delve and they just say, okay, you’re saying you’re fine.” [I13]
- “I would still be rather hesitant in fully trusting that they understand what my symptoms are and that they’re giving me the right advice. Because most of the advice would be based on what I’m telling them and I’m not a clinician.” [I15]

*Subtheme 1.1.2: Lack of formal guidance*

- “We’re operating without any direction as to ‘this is safe’ or ‘this is correct or not’. We’re just using our core clinical judgement.” [FG1 P5]
- “There are divergent views within my practice as to the threshold as to what requires a face-to-face consultation and what can be done remotely…I’m not aware of any formal guidance” [I25]

*Subtheme 1.1.3: Defensive medicine*

- “There is a sort of issue that GPs might like to practise defensive medicine [and] issue prescriptions inappropriately, because they don’t know the severity of it” [FG3 P4]
- “I think you have to have a lower threshold, to sending [patients] to hospital. Probably more threshold than you would when you actually see them face-to-face.” [I9]

###### Theme 2. Negative Impact on Patients’ Access to Care

*Subtheme 1.2.1: Delays in care*

- “I’ve noticed that [it] is difficult if you don’t get much information from a patient…then you do a telephone consultation. And then say, Well, we need a video consultation, and then a video consultation, and then a face-to-face. So one of the challenges is not getting enough information in the first point of contact because then it delays things.” [FG1 P5]
- “…[I am often] astonished when you’re told it’s a two week wait, and what you’re requesting is some urgent medication or referrals of a post-operative care…” [FG1 P4]

*Subtheme 1.2.2: Poor continuity*

- “I was getting loads of treatment, but nothing that worked until my fifth consultation, which ended up as a visit and then it was solved like that…I felt because it was phone calls, and I was constantly speaking to different people, I felt like a nuisance. So it was really something that could have been solved in one 10-minute visit that took three weeks....You should be able to say, I saw Doctor X; I need to speak to Doctor X again rather than have to go through the whole thing again with Doctor C. So again, it’s continuity and having that point of contact.” [I28]
- “The other difficulty [in virtual care] is getting to speak or to see the same doctor more than once. And that I think is a major, major problem in large practices. You get a lot from a patient by seeing them at different intervals and seeing what’s changed over a long period of time…And on the phone, you get absolutely nothing of that and by seeing a different doctor, every time you get nothing of that either.” [FG3 P1]

*Subtheme 1.2.3: Digital exclusion*

- “In the Northeast, we have the lowest rate of computers that people have access to in their own homes…My concern is that the discrepancy between those who can afford broadband, and who can afford to have access to a laptop, iPad, whatever, at home, is going to get wider; it’s not narrowing. ” [FG1 P4]
- “So, not always, but certain elderly people that either don’t know how to use their device or having problems with hearing, or people that have dementia for example and are trying to have a consultation remotely, even if there’s another person present, is even more challenging than it is in person.” [I2]
- “Minority populations where women may not speak the language…And so, access to a GP or medical help is through, you know, family members, sometimes the husband or other family members. I felt uncomfortable before, where I wasn’t sure if I was getting all the information.” [I9]

*Subtheme 1.2.4: Patients’ use of alternative routes/services to access health care*

- “So what I envisaged that the safety problems have been is that people ended up going to A&E or calling 111, and that delays the process of being seen.” [FG2 P5]
- “They fear that this is just a back door way of saying yes we are doing healthcare, but they don’t really care about me as an individual until I’m on my deathbed. And I say that because again and again, I’m hearing one thing and one thing only, that I cannot get an appointment to go and see a GP. So, most people have now effectively switched off and they’re not even trying to get a GP appointment. They try to paddle along on a daily basis until their health deteriorates so much that they have to call an ambulance or they end up going into A&E and then they’re hospitalised.” [I15]

###### Theme 3. Worsening the Workforce Crisis

*Subtheme 1.3.1: Digital fatigue*

- “It’s just another route from which patients can consult and contact the practice and already we’re having to manage emails and manage our kind of record system with tasks and notes coming through that or getting messages now through our text messaging service and photos as well through that. Getting letters. You know, we’re getting all sorts of things in lots of different directions and it just is overwhelming” [I25]
- “The problem with all these tools and everything like that, is it actually it just adds more work (…) at the moment, when I go in, I have to monitor emails, tasks, e-consultation messages, instant messages. (…) So there are lots of modalities of accessing the GP, you get messages all over from, and then you get someone phoning up. (…) So another tool or another platform, another password, no thank you.” [I4]

*Subtheme 1.3.2: Decreased worker satisfaction*

- “I personally think it’s done harm to the image of General Practice. Because people feel they are being fobbed off because doctors won’t see them. You only have to read the comments (…) saying ‘GPs are lazy’, ‘don’t see patients’, ‘tell everyone to go to A&E’, ‘why don’t they see patients anymore?’. And part of that is because the work is hidden away now because you’re like a call centre.” [I4]
- “Sometimes I think when you’re doing a lot of remote triage, this is not what I thought that my role would be. You know, like, when I imagined myself as a doctor, I didn’t really think I was going to be spending so much time on the phone. So it’s like almost, I dare say, professional identity crisis, but it is very different from what I thought I would be doing.” [I9]

### Strategies to Mitigate Patient Safety Risks in Virtual Primary Care

Despite the number of risks identified by both patients and providers, both groups put forth both overlapping and mutually exclusive suggestions for improvement. An overview of these potential solutions is provided in Figure 1.

**Figure 1.**
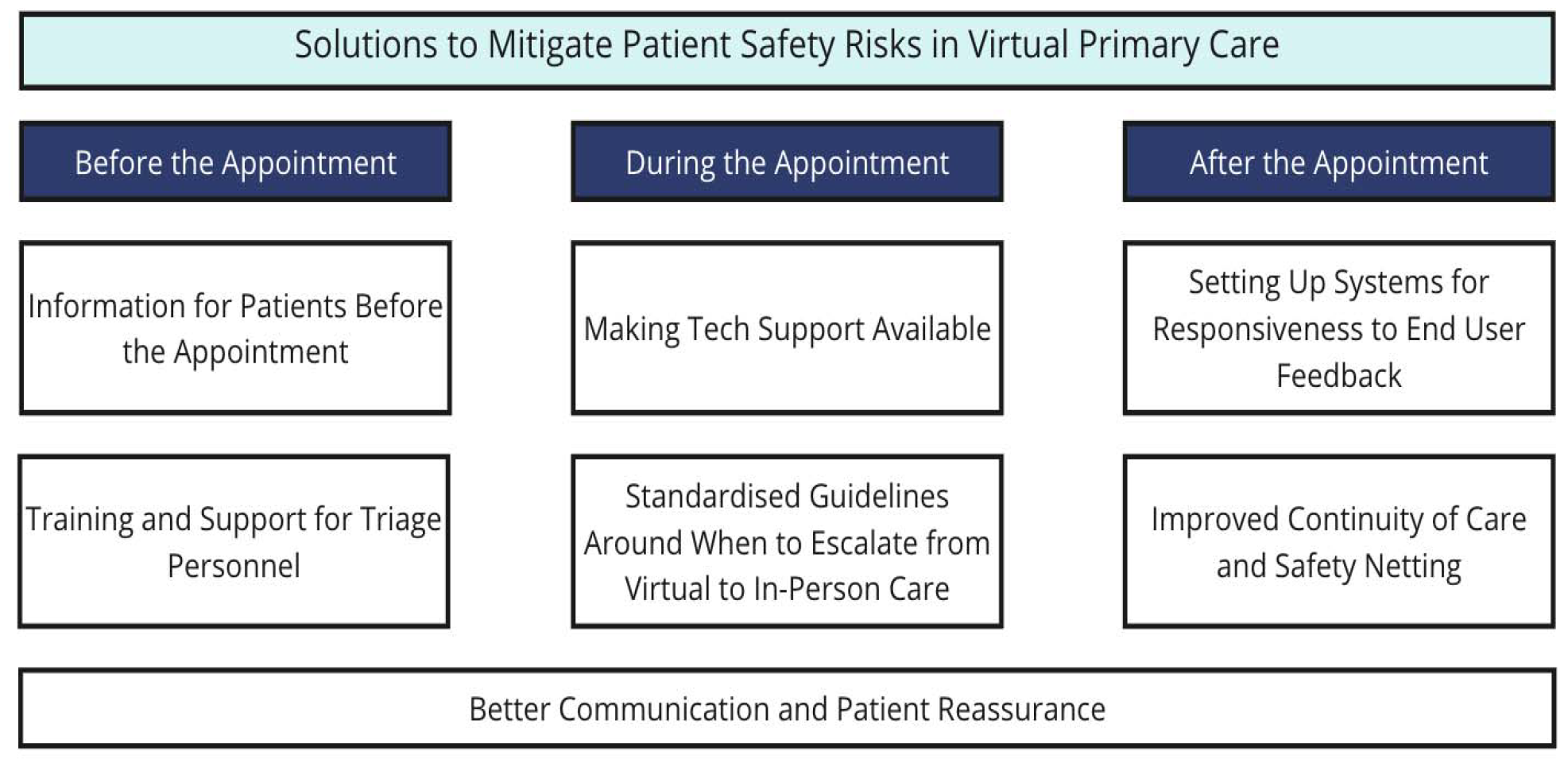
Overview of strategies identified by patients and providers.

Both patients and providers noted that there is value in equipping the patient with information before a virtual appointment, to better direct patients to the appropriate service. Participants indicated that the NHS website itself may be best suited as the venue for this information, with the caveat that it be redesigned for improved user experience. Providers perceived value in equipping patients with ways to ‘collect’ information needed for an effective clinical assessment (e.g., digital) but emphasised the importance of patient education in this context. Both providers and patients emphasised the role of receptionists and triage personnel in acquiring important information before the appointment to inform the choice of the appropriate modality of consultation, often citing the importance of training to do so.

Technology challenges during virtual care were described as burdensome and distracting. Participants recognised the importance of establishing ‘backup plans’ for instances in which the connection was lost. Improved technological support was prioritised, with participants citing the need for readily available IT expertise to troubleshoot the issue. With a significant, but not exclusive, emphasis on the virtual care technology, participants described the importance of systems for feedback to developers about the virtual care experience. Participants indicated that visible responsiveness to feedback and regular updates on changes in technologies would likely increase their willingness to use the platforms for virtual care delivery.

Improved continuity of care was deemed important by participants and its recommended operationalisation took a number of forms. One participant highlighted the increased importance of (and burden on) caregivers in maintaining continuity of care in the virtual environment, particularly when escalating from virtual to in-person care.

A cross-cutting theme across each step of the virtual care journey was the importance of reassurance and communication with patients. The onus was placed on the providers to ensure their communication was not perceived as transactional and that the relational needs of patients were still met virtually. Participants illustrated examples of mutual agreements between the patient and provider around what to monitor after the virtual appointment and what to do if concerned; this form of virtual care safety netting provided participants with a sense of confidence in the next steps. One participant mentioned the utility of reassuring patients that the information they were receiving was from a validated source.

Altogether, participants suggested that these strategies can collectively contribute to reducing existing inequities in accessing primary care.

#### Textbox 2

**Strategies to Mitigate Patient Safety Risks in Virtual Primary Care**

##### Theme 2.1: Information for patients before the appointment

- “I just sent them a link sometimes about what to expect and how to prepare for the video, like use good lighting, find a quiet room, so there are no distractions…Having a very clear agenda beforehand about what the video consultation is going to be about is really useful. And it might help you to plan how you’re going to manage the situation because if they’ve got multiple problems, it may have just been quicker just to bring them in for a face-to-face from the start.” [I3]
- “[The website] may even help them think whether the service that they’re coming into is the correct one for them. So say they had features of cauda equina syndrome, they should probably be in A&E.” [I3]
- “There needs to be… some kind of facilities to complement the video consultation. One of the key things we found when we tried to do that was doing it in real-time is not very efficient. So trying to get someone to use a digital stethoscope for the first time, once you’re in the middle of a video consultation is not good.” [I3]

##### Theme 2.2: Training and support for triage personnel

- “If the receptionists are well trained, they can signpost quite well as to whether this needs to go immediately to a more urgent treatment centre or whether they’ll be able to wait for a telephone call later that day.” [FG1 P1]
- “I would like there to be a robust system for the screening. So instead of having the initial call with a receptionist, and you have it with the dedicated healthcare assistant or a nurse, because then if they can filter you to where you’re supposed to be going…Because if you go through, say a receptionist because they’re not medically trained, then you might just be actually duplicating work and wasting time” [FG3 P2]

##### Theme 2.3: Making tech support available

- “Who to step things up to when there’s a problem. So, if there’s a technical problem, is there a person that I can then reach out to if there’s some other safeguarding or whatever other issue, where do I go with that?” [I2]
- “The idea of an IT-type person, an IT or digital person at the practice, would probably be good. Someone that is physically there that you can go to when you’re having some kind of issue and that they’re specifically trained on that system and with those issues to help you.” [I2]
- “Laying out [the next steps if there’s a problem], for example, if something does go wrong, and there’s a loss of connection” [I27]

##### Theme 2.4: Standardised guidelines around when to escalate from virtual to in person care

- “Just to be more aware of this threshold when you’re worried that a patient needs to be seen or, or in hospital…I guess if you go through examples in training, for example, that helps a little bit” [I9]
- “ I think not having an obvious place to access those guidelines and referral requirements really slows down patient care. And you often don’t find out about it until a couple of weeks down the line when your referral is returned, or the triage summary is sent back to you with this, that and the other. So that would be really helpful if we had a clear access to … really local guidelines, not national guidelines.” [I24]
- “It’d be useful to have things like scoring systems for remote care, such as the fever pain scores, for when …you can’t examine the tonsils….Would be really helpful to give us that reassurance that okay, am I doing the right thing or not because there is so much uncertainty.” [FG1 P5]

##### Theme 2.5: Setting up systems for responsiveness to end-user feedback

- “Depending on who’s made the program, they might feel that it’s an intuitive thing, but I don’t think it always is, and I’d like to see some feedback going back the other way when things aren’t working…They have a feedback form, where you can say what’s going on and then visibly within a couple of months you see that something is happening, and they keep you updated on the changes.” [I2]
- “Checking in with people, do you feel comfortable, do you like the system, don’t you like the system, why don’t you like the system, both from the staff side and the patient side, because if people don’t like using something, they’re not going to use it, or they’re not going to use it well and that could also possibly cause patient safety issues.” [I2]
- “We need to get that trust…Don’t expect the communities to come to you. We need to go out there into the communities, need to try to understand the challenges that these communities are experiencing on a daily basis. Whether it’s to do with simple, basic healthcare needs or whether it is about literacy of healthcare needs.” [I15]

##### Theme 2.6: Improved continuity of care and safety netting

- “Sort of guidance for carers in that sort of instance. You know, this is what you should be looking out for. Please get in touch if this is the case or if this isn’t the case then have to do that.” [I25]
- “It can be a bit ad hoc as to which ones are sent…It’d be really nice if there was a suite of resources there of safety netting materials that were really high quality.” [I3]
- “Do you feel you would benefit from seeing me face-to-face or are you happy with what I’ve suggested to try out for a couple of weeks and if it doesn’t work out then you come back and see me. So all those kinds of reassurances I think would be good to finish off.” [I12]
- “I suppose maybe like some sort of a feedback survey… It’s almost like that flowchart sort of system working, Is your condition more under control? If you said “no”, then you’d automatically be prompted then to rebook in… So…no one’s falling behind. ” [I17]

##### Theme 2.7: Better communication and patient reassurance

- “It’s giving them that level of confidence, engaging with them, understanding the routes, and then giving them reassurance that actually it is the human being who will be interacting with, you’re not talking to the computer.” [I15]
- “I don’t think most GPs change their communication skills significantly even though it is a different format or if they do it’s just more closed questions and closed questions can lead to a transactional approach. So, training on how to get that relational care through remote consulting would be useful.” [I25]

## Discussion

### Summary of Key Findings

Three main areas for patient safety risks associated with virtual primary care were identified, including suboptimal clinical decision-making, negative impact on patients’ access to care, and worsening the workforce crisis. Strategies to mitigate these risks included providing information for patients, training triage personnel, making technical support available, standardising guidelines, setting up systems for feedback, improving continuity of care, communication, and safety netting.

### Findings as Compared with Previous Studies

The challenges of obtaining meaningful patient information and conducting accurate virtual assessments are congruent with previous literature. A systematic review by Ftouni et al., reported concerns of compromised physical assessment capabilities in the virtual care setting (16), and additional evidence suggests practitioners experience more uncertainty in virtual clinical decisions due to the compromised ability to conduct a reliable clinical assessment (7, 17). This study supports these findings and adds that there is an increased demand placed on the patients to take on the role as a ‘supplier’ of the information needed for clinical decision-making.

In addition to the challenges of conducting a clinical assessment in virtual care, there is also a unique barrier to establishing a holistic understanding of the patient’s health and well-being due to the compromised ability to perceive non-verbal cues. Previous studies have suggested that providers perceive virtual care consultations to be less information-dense for these reasons (7, 18). Our study largely corroborates these findings and highlights how other pre-existing in-person challenges (e.g., language barriers) can be further exacerbated.

Our findings corroborate unique virtual care access challenges based on a range of disadvantages, such as older age or access to technologies. Even before the marked increase in the use of virtual care technologies, lack of access to care has been a significant contributor to the underutilisation of preventive services and therefore poorer outcomes (21). Though some of these barriers have been attenuated with the introduction of virtual care (22), virtual care has introduced another layer of care access challenges that particularly impact communities that are often already disadvantaged. Previous research exploring the virtual care access challenges has found that increased use of virtual care consultations may exacerbate the digital divide for vulnerable patients (23-25).

In line with the findings of this work, previous literature has also highlighted that virtual care can decrease provider job satisfaction (7). Specifically, both patients and providers have cited concerns of the worsened providers’ work-life balance with increased use of virtual care (26).

### Strengths, Limitations, and Future Research

This study used a topic guide developed based on existing literature and findings about remote care from earlier studies undertaken by our research group, and engaged both practitioners and patients, stimulating particularly rich discussions. The subsequent analyses were performed by two independent researchers with backgrounds in clinical medicine, public health, and patient safety, in consultation with other senior researchers with further expertise in qualitative research methods. The focus groups were conducted online which helped to maximise its accessibility to interested participants located all across the UK.

Nonetheless, some limitations need to be acknowledged. While our study was focused on the usage of virtual care tools in primary care as a whole, what this meant for participants varied greatly. The broad definition used during the focus groups and interviews limited our ability to form a deeper understanding of and recommendations for specific forms of virtual care technologies. However, this could have been potentially offset by the fact that the diverse range of views captured simply reflected the myriad of virtual care tools currently in use across the NHS. Though patients were recruited from diverse sources using a variety of different approaches, recruitment was limited to the external platforms accessible to the research team; hence, some groups may have been missed (e.g., those not engaged with online patient communities). Despite prioritising diversity in our recruitment, the majority of the participants were white females. Thus, our study might not have sufficiently captured the experiences of practitioners or patients from ethnic minority backgrounds.

It is important to note that this study examined subjective perceptions. Though these findings are important in exploratory, in-depth research, it is imperative to expand on these qualitative findings with additional objective measures of patient safety to understand if, how and to what extent the perceived safety risks come to fruition in virtual care settings.

As illustrated in this study, the safe use of virtual care is contingent on patients’ availability of the necessary technology, their knowledge of how to use technology to meaningfully engage, and their perceptions of privacy. Further research is needed on how to minimise the digital divide and acknowledge unique nuances for different populations. This may include follow-up studies focusing on individuals with language barriers, lack of stable addresses, low digital skills, or complex needs.

### Implications for Policy and Practice

Both patients and providers experienced uncertainty around when the use of virtual care is or is not suitable, highlighting the need for clear guidance and policies to minimise uncertainty in triaging for virtual care delivery. Policies that promote greater collaboration between virtual care and face-to-face settings can encourage a holistic representation of a patient’s journey and enhance their overall experience when seeking care in the health system. For example, if an influx of patients with respiratory symptoms is anticipated for flu season during the autumn months, it would likely be useful to develop standardised guidance clearly delineating how patients should access care from virtual care and in-person consultations, while maintaining flexibility for adaptation to ongoing and unforeseen safety challenges (28).

Addressing the perception surrounding the supposed ‘inferiority’ of virtual consultations would likewise need to be multifaceted. This may range from indirect measures such as improving how novel technologies are systematically implemented into clinical settings, supporting more robust technical support, and providing a robust evaluation of the impact of the different modalities on the quality of care delivered. Introducing better guidance on how to standardise the safety netting discussions at the end of virtual care appointments can also contribute to mitigating some of the perceptions that virtual consultations are ‘inferior to’ in-person consultations.

Given that providers cited significant challenges and frustrations with using subpar technologies for virtual care visits, standards for a thorough evaluation of any new technology should be applied to ensure the technology, if adopted, is congruent with the existing clinical workflows. Continuous communication between virtual care technology developers and end users is crucial to ensure end-user satisfaction. A continuous feedback loop between end-users and developers can provide real-world insights, ensuring that it reflects the end-users’ needs and preferences.

## Conclusion

The uptick in the use of virtual care delivery during the COVID-19 pandemic has demonstrated the complex and often intertwined relationships between patient safety risks, benefits, challenges, and potential strategies. The pandemic has expedited the use of these technologies and made apparent the need for improved and better-coordinated guidance surrounding their use. Our study has highlighted that patients and providers now have a heightened awareness of the strengths and pitfalls of virtual care delivery, due to their increased familiarity with the use of virtual care technologies. Given the greater clarity around how this technology can support care delivery, existing policies need to be updated and new policies need to be devised to minimise risks associated with virtual care and support patient and provider workflows.

## Data Availability

Data is available upon reasonable request to the corresponding author.

## Acknowledgements

OL, EL, JvD, NB, AA, and ALN contributed to the conception and design of the study. All authors contributed to writing the manuscript, provided critical revision and approved the final version of the manuscript. OL, EL, and ALN guarantee the integrity of the work.

## Declarations

### Consent for Publication

All participants provided written informed consent for anonymised quotes from their interviews to be used in the publication of this study.

## Availability of Data and Materials

Data is available upon reasonable request to the corresponding author.

## Appendices

**Appendix 1** – Focus group topic guide.

**Appendix 2** – Semi-structured interview topic guide.

**Appendix 3** – Study participant consent form.

