## Appendix 1 for "Patient Safety of Remote Primary Care: A Qualitative Study Assessing Risks and Mitigation and Prevention Strategies"

**Safety Checklist Remote Consultations - Topic Guide**

**Facilitator roles**

Facilitator 1: Read the questions, chair the discussion, and monitor hands up (either using zoom functionality or video). Ensure anyone joining by phone has opportunity to contribute to all discussions.

Facilitator 2: Manage the technology e.g. admit people from waiting room, divide participants in breakout room (only relevant for focus group on 1 Dec); help with any technological difficulties. Also monitor chat and input comments from there into discussion (using hands up function so that facilitator 1 can see when to invite incorporation of chat into the discussion). Facilitator 2 may need to join from two devices – one to record, and one to be able to type into and do screen sharing.

**Introduction to the focus group (5-10 minutes)**

1. Give introduction of team
2. Ground rules using Zoom: ‘hands up’, use of chat, not referring to names of people or organisations
3. Confirm that the interviewees understand:

- The purpose of the research
- What the focus group or interview entails
- How confidentiality and anonymity will be assured
- That they can stop at any time without explanation

And that:

- They have had the chance to ask questions
- They are content to be recorded and that they have the option to switch off their video and use only first names on their Zoom square if they wish

4. Definitions
   - ***Definition of ‘remote consultations’ (Slide 1)***
   - ***Definition of ‘patient safety’ (Slide 2)***
   - ***Pathway of care (Slide 3)***
5. Handover to facilitator 2 to divide into breakout rooms and/or begin recording

**Introductions to each other (10 minutes)**

1. Introductions of participants to each other (5 min)

**Main body (40 minutes)**

***Online triage + remote consultations together (45 minutes)***

1. When trying to access or deliver GP consultations during the pandemic, what technologies did you use?
2. Based on your experiences of receiving, giving or observing online triage and/or remote consultations, tell us about any times when something went wrong or nearly went wrong, or when you felt the care was unsafe.

- Based on your experiences and concerns, what kind of support would help **patients and/or healthcare professionals** to have safe remote consultations?
  - Probe: What would such a source of support look like (e.g., training, checklist, information sheet etc)?
  - Probe: How and when would you like to receive this support (e.g. before or during the consult? Via text message, e-mail, flyers etc?)
  - Probe: What might be barriers or facilitators in using such resources in practice?

**Closing (1-2 minutes)**

- Anything else anyone would like to add?
- Thank participants
- Next steps for reimbursement: NPF forms will be shared for completion
