## Appendix 2 for "Patient Safety of Remote Primary Care: A Qualitative Study Assessing Risks and Mitigation and Prevention Strategies"

**Project title: Evaluating and improving remote consultations in primary care (SEMI-STRUCTURED INTERVIEW TOPIC GUIDE)**

**Going over the Participant Information Sheet:**

- Overview and purpose of study
- Aims of the focus and expected duration
- Why participant’s involvement is important, advantages and disadvantages
- What will happen to the results of this study?
- Any questions before we begin the session?

| **Questions for GPs /HCPs** | **Questions for patients** |
| --- | --- |
| In your opinion, what makes for a safe virtual consultation? | In your opinion, what makes for a safe virtual consultation? |
| Were there any moments where you ever felt/experienced that the use of virtual consultations was unsafe? Can you provide any examples? | Were there any moments where you ever felt/experienced that the use of virtual consultations was unsafe? Can you provide any examples? |
| We would like you to think about what kind of organisational support do you think would help to better manage virtual consultations.   - What you like to have support to better manage the technical aspects (hardware, software)? If so, which support? - What you like to have support to make safer decision making? Is so, which support (i.e., training, guidelines, decision support tools)? - Are there other support aspects that you’d like to see considered? | We would like you to think about what kind of support could healthcare organisations give you, so that you would better manage virtual consultations.  *“Getting a consultation appointment”*   - **Awareness of remote consultations:** Are patients aware of remote consultations? What can we do to improve this? (e.g., GPs promoting it in person prior to first remote session, pamphlets, social media, community outreach etc.,) - **Determining whether remote consultations are appropriate** for patient’s presenting complaint vs. F2F: Do patients find that they need help figuring out whether remote consultations are right for their health problems? If so, how can we address this dilemma? - **Access to remote consultations:** Are there any changes which would better help you get access and book remote consultations? (e.g., infrastructure, how to use tutorials, interoperability between HCPs/facilities frequented)   *“The consultation itself”*   - What would make the **remote consultation experience itself,** ‘better’ for patients? How could we achieve that from patient/GP/NHS perspectives? (e.g., importance of video, confidentiality, HCP communication skills, etc.,) - What can be changed in the remote consultation session itself which would **make it safer** for patients? - What changes would **make patients want to continue to use remote consultations** moving forward?   *“After the consultation”*   - How can patients be supported in their next steps after the virtual consultation?   **Support could include training, written guides, virtual support, improved user interface, etc.* |
| Would you like to have any specific training on remote consultations?   - If yes, what **clinical topics** would you like to see covered as part of your training on remote consultations? *(Prompts: carrying an effective clinical consultation, risk stratification & prioritisation, manage clinical risk, guidance on how to perform remote consultations)* - Thinking specifically about training programmes, what **non-clinical topics** would you like to see covered as part of your training on remote consultations? *(Prompts if needed: support patients that struggle with technology, technical skills, patient confidentiality, information governance, remote communication skills)* - Thinking about training in remote consultations, at which level do you think it should be delivered (undergraduate, during medical speciality, later)? - How would you prefer this training to be delivered (i.e., written materials, e-learning, webinars, face to face, peer support, direct observation, case bases discussion?) | Would you like to have any specific training on remote consultations?   - If yes, are there any topics that you would like to see covered? - How would you prefer this training to be delivered (i.e., written materials, e-learning, webinars, face to face, peer support?) |

**Closing: Is there anything else that you think is important about using remote care tools, virtual consultations, and patient safety that we have not talked about?**

- Summarize covered research domains/questions.
- Any other questions or concerns?
- Ensure participant has copies of participant information sheet and consent forms; provide any additional information as needed
- Thank participants for their input and engagement in the discussion.
