## Appendix 3 for "Patient Safety of Remote Primary Care: A Qualitative Study Assessing Risks and Mitigation and Prevention Strategies"

### Consent Form for Participants Able to Give Consent

**Full Title of Project: A safety checklist for remote consultations in primary care**

**Name of Principal Investigator:** Dr Ana Luisa Neves, Associate Director / Advanced Research Fellow, NIHR Patient Safety Translational Research Centre, Imperial College London

**Co-investigators**: Jackie van Dael; Edmond Li; Monsey McLeod; Alay Rangel (Imperial College London)

**Please initial box**

| 1. I confirm that I have read and understand the participant information sheet version 1.0 dated 23/04/2021 for the above study and have had the opportunity to ask questions which have been answered fully. |
| --- |
| 2. I understand that my participation is voluntary, and I am free to withdraw at any time, without giving any reason and without my legal rights being affected. |
| 3. I give permission for Imperial College London to access my research records that are relevant to this research. |
| 4. I give/do not give (delete as applicable) consent for information collected about me to be used to support other research in the future, including those outside of the European Economic Area (EEA). |
| 5. I consent to take part in the above study. |
| 6. I give consent to my participation in a video interview being recorded. |
| 7. I give consent to being contacted about the results of the study. |

________________________	________________	________________

Name of participant	Signature	Date

_________________________	________________	________________

Name of person taking consent	Signature	Date

(if different from Principal Investigator)

_______________________	________________	________________

Principal Investigator	Signature	Date

1 copy for participant; 1 copy for Principal Investigator
